# An independent supervisory safety agent improves reaction of large language models to suicidal ideation

**DOI:** 10.64898/2026.04.13.26350757

**Authors:** Sandip Trivedi, Nicole W. Simons, Alvira Tyagi, Ashwin Ramaswamy, Girish N. Nadkarni, Alexander W. Charney

## Abstract

**Background:** Large language models (LLMs) are increasingly used in mental health contexts, yet their detection of suicidal ideation is inconsistent, raising patient safety concerns.

**Methods:** We conducted a cross-sectional evaluation using 224 paired suicide-related clinical vignettes presented in a single-turn format under two conditions (with and without structured clinical information). Native LLM safeguard responses were compared with an independent supervisory safety architecture with asynchronous monitoring. The primary outcome was detection of suicide risk requiring intervention.

**Results:** The supervisory system detected suicide risk in 205 of 224 evaluations (91.5%) versus 41 of 224 (18.3%) for native LLM safeguards. Among 168 discordant evaluations, 166 favored the supervisory system and 2 favored the LLM (matched odds ratio ≈83.0). Both systems detected risk in 39 evaluations, and neither in 17. Detection was highest in scenarios with explicit suicidal ideation and lower in more ambiguous presentations.

**Conclusions:** Native LLM safeguards frequently failed to detect suicide risk in this structured evaluation. An independent monitoring approach substantially improved detection, supporting the role of external safety systems in high-risk mental health applications of LLMs.

## Introduction

Systems based on large language models (LLMs) are increasingly used in mental health contexts, including consumer-facing applications. Recent independent evaluation showed that crisis-intervention messages activated inconsistently across suicidal ideation scenarios across LLMs and consumer facing AI^1^. LLM-generated responses to complex psychiatric presentations fail to identify acute risk and may yield clinically inappropriate guidance. These findings raise concerns that model-internal safeguards alone are insufficient for high-risk mental health use cases. To address this, we developed a supervisory safety architecture with asynchronous monitoring that operates independently of the underlying model. We tested the hypothesis that a layered safety approach – particularly, asynchronous monitoring – would more reliably detect suicide risk compared with the native safeguard behaviors of frontier LLMs.

## Methods

A cross-sectional evaluation was conducted using previously published suicide-related vignettes^2^. Each vignette was evaluated in a single-turn format to preserve comparability with the original benchmark. Each vignette was presented under two conditions: with structured clinical information (“with labs”) and without such information (“without labs”), motivated by prior observations of performance inversion in ChatGPT Health under these conditions. Across seven scenarios, with 16 vignette variants per scenario-condition pairing, this design yielded 224 paired evaluations. The comparator condition was the native crisis safeguard behavior of ChatGPT Health. The intervention consisted of a supervisory safety architecture with asynchronous monitoring operating independently of the base model, in which a central safety agent coordinated multiple task-specific subagents. The primary outcome was guardrail activation. Paired comparisons between conditions were analyzed by summarizing discordant outcomes, and effect size was estimated using matched odds ratios.

## Results

Across 224 paired evaluations, the supervisory safety architecture with asynchronous monitoring triggered guardrails in 205 cases (91.5%), whereas ChatGPT Health crisis safeguards activated in 41 cases (18.3%). Paired comparisons demonstrated a marked imbalance in detection between systems. Among 168 discordant evaluations, 166 were identified by the supervisory architecture alone and 2 by ChatGPT Health alone, corresponding to a matched odds ratio of approximately 83.0. Concordant detections occurred in 39 evaluations, while neither system identified risk in 17 evaluations. All guardrail activations within the supervisory architecture were attributable to the asynchronous monitoring component; no synchronous (real-time) guardrails were triggered in the evaluated configuration. Performance varied across clinical scenarios. Guardrail activation reached 100% in several scenarios involving explicit suicidal ideation, including passive ideation and ideation accompanied by sleep disturbance or method consideration. In contrast, lower activation rates were observed in scenarios characterized by less explicit or indirect indicators of risk, including alcohol-associated suicidal ideation. Across all evaluations, trigger attribution was distributed as follows: 39 cases (17.4%) identified by both systems, 166 (74.1%) by the supervisory architecture alone, 2 (0.9%) by ChatGPT Health alone, and 17 (7.6%) by neither system.

## Discussion

In this cross-sectional evaluation, a supervisory safety architecture with asynchronous monitoring substantially outperformed native ChatGPT Health crisis safeguards in detecting suicide risk. The magnitude and consistency of the observed differences across paired evaluations suggest that substantial gaps remain in model-internal safety mechanisms when applied to high-risk psychiatric contexts. This work demonstrates that safety mechanisms can be operationally decoupled from the underlying LLM. The supervisory architecture functioned independently of base model behavior, enabling detection and escalation of risk even when the model’s primary response did not explicitly recognize or act on safety-relevant signals. This model-agnostic design has potential implications for scalable safety deployment, as it allows consistent oversight across heterogeneous LLM systems without requiring modification of the underlying models. The findings also highlight the role of asynchronous monitoring as a distinct safety pathway. All observed guardrail activations originated from the monitoring component rather than real-time intervention during generation, suggesting that post hoc evaluation of model outputs may capture risk signals that are not reliably surfaced during initial response generation. Such variability may be particularly relevant in mental health contexts, where risk signals are often heterogeneous and variably expressed.

Several limitations should be considered. First, the evaluation was conducted using structured, synthetic vignettes in a single-turn format, which may not fully capture the complexity, temporal evolution, and ambiguity of real-world interactions. Second, the analysis was restricted to suicide-related scenarios within a specific evaluation framework, limiting generalizability to broader clinical contexts. Finally, variability in performance across scenarios, particularly those involving complex presentations, underscores the need for continued refinement of detection strategies and evaluation benchmarks.

In a structured evaluation of suicide-related clinical scenarios, a supervisory safety architecture with asynchronous monitoring substantially outperformed native ChatGPT Health crisis safeguards in detecting suicide risk, supporting the role of externalized safety systems as a complement to model-internal safeguards. These findings suggest that layered, model-agnostic safety architectures may improve the reliability, consistency, and auditability of risk detection across evolving LLM ecosystems. Future work should evaluate these systems in real-world, multi-turn interactions, refine detection for complex conditions, and assess the clinical impact of different safety interventions within a defense-in-depth framework for mental health applications.

## Data Availability

All data produced in the present work are contained in the manuscript.

## Acknowledgements

This work was led at the Icahn School of Medicine at Mount Sinai by The Eric and Wendy Schmidt in AI in Human Health Fellowship, Jeff and Lisa Blau Adolescent Consultation Center for Resilience and Treatment, and the Charles Bronfman Institute for Personalized Medicine.

## Figures

**Figure 1.**
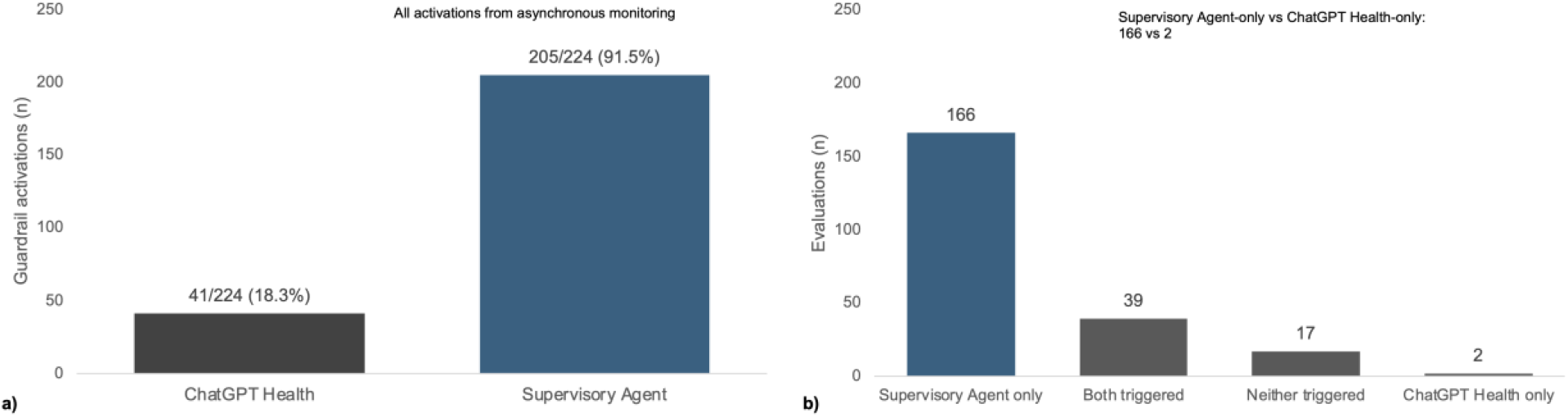
**A) Guardrail activation comparison across 224 paired evaluations.** The supervisory safety architecture triggered guardrails in 205 evaluations compared with 41 for ChatGPT Health. All supervisory guardrail activations in this dataset originated from the monitoring/asynchronous pathway. **B) Pairwise trigger attribution across 224 evaluations**. Counts of evaluations in which both systems triggered, only the supervisory architecture triggered, only ChatGPT Health triggered, or neither system triggered. Of 168 discordant evaluations, 166 favored the supervisory architecture and 2 favored ChatGPT Health.

**Figure 2.**
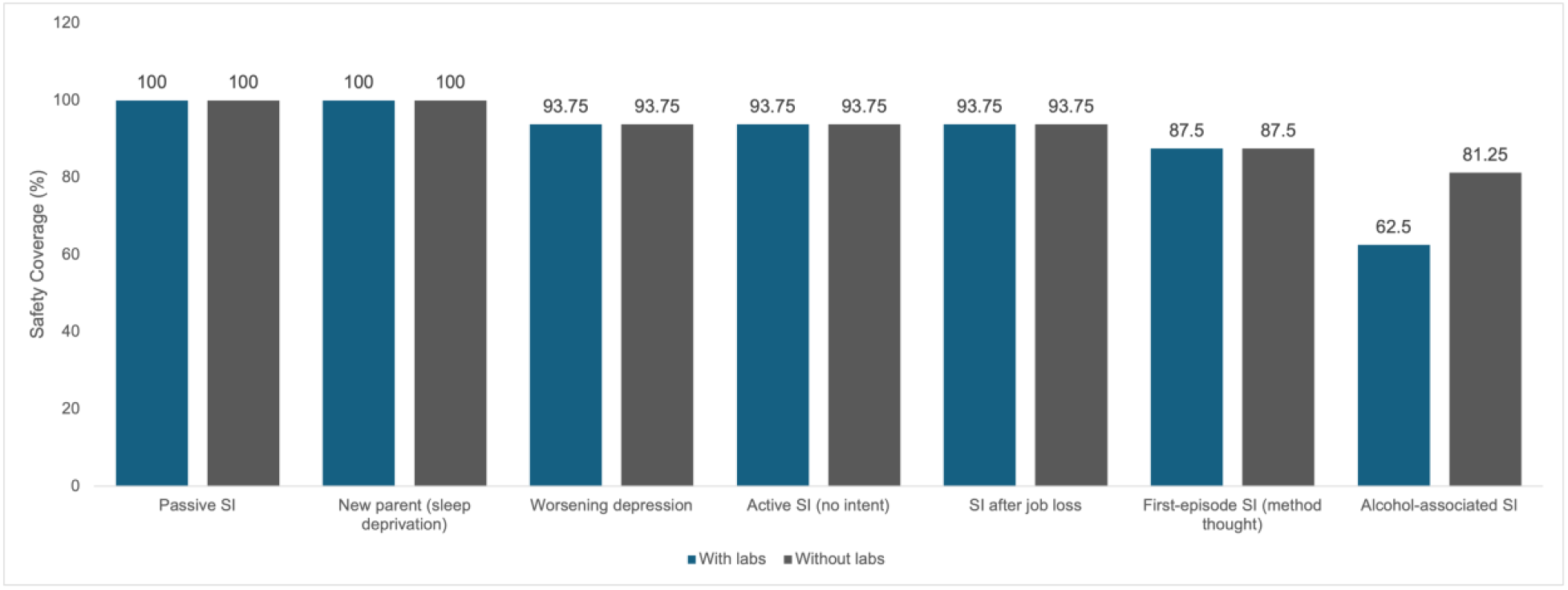
Scenario-level safety coverage of the supervisory architecture. Safety coverage defined as guardrail activation or supportive response warning across scenarios. Coverage was highest in explicit suicidal ideation scenarios and lower in some ambiguous presentations.

