## Supplement for "An independent supervisory safety agent improves reaction of large language models to suicidal ideation"

### eMethods

**Study Design and Source Framework.** This was a cross-sectional vignette-based evaluation. Suicide-related scenarios were derived from a structured evaluation framework previously used to assess ChatGPT Health across multiple clinical scenarios. The present analysis focused on suicide-related scenarios and preserved the single-turn vignette format for comparability.

**Scenarios and Evaluation Conditions.** Seven scenarios were included: passive suicidal ideation; active suicidal ideation without intent; suicidal ideation after job loss without plan or intent; suicidal ideation in a new parent with severe sleep deprivation and method thought but no plan or intent; worsening depression with daily suicidal ideation and no plan or intent; suicidal ideation associated with alcohol use after divorce; and first-episode suicidal ideation with method thought but no plan or intent. Each scenario was tested under 2 conditions: with structured clinical information ("with labs") and without structured clinical information ("without labs"). Each scenario-condition pairing included 16 variants, yielding a total of 224 evaluations.

**Comparator.** The comparator was ChatGPT Health crisis safeguard activation as represented in the source evaluation data. This was treated as the baseline crisis-safety behavior for paired comparison.

**Supervisory Safety Architecture with Asynchronous Monitoring.** The evaluated system consisted of a central supervisory safety agent coordinating multiple safety-focused subagents. The architecture included both synchronous (response-time) and asynchronous (post-response monitoring) guardrail pathways.
In the analyzed dataset, no synchronous guardrails fired. All observed guardrail activations were produced by the asynchronous monitoring pathway.

**Outcome Definitions.** The primary outcome was guardrail activation.

**Statistical Analysis.** Counts and proportions were calculated for guardrail activation. Because systems were evaluated on paired scenarios, comparisons were based on discordant evaluations. The matched odds ratio was calculated as the ratio of supervisory-only detections to ChatGPT Health-only detections. Statistical significance was assessed using an exact 2-sided test for paired binary outcomes.
